# Factors associated with drinking behaviour during COVID-19 social distancing and lockdown among adults in the UK

**DOI:** 10.1101/2020.09.22.20199430

**Authors:** Claire Garnett, Sarah Jackson, Melissa Oldham, Jamie Brown, Andrew Steptoe, Daisy Fancourt

## Abstract

**Aim:** To assess what factors were associated with reported changes to usual alcohol drinking behaviour during the start of lockdown in the UK.

**Design:** Online cross-sectional survey from 21^st^ March to 4^th^ April 2020.

**Setting:** UK.

**Participants:** 30,375 adults aged ≥ 18y.

**Measurements:** Changes in drinking over the past week, sociodemographic characteristics, diagnosed or suspected COVID-19, adherence to COVID-19 protective behaviours, stress about COVID-19, finances or boredom, recent drop in household income, key worker status, and health conditions.

**Findings:** Of 22,113 drinkers (65.7% of analytic sample), 48.1% (95% CI=47.0-49.1%) reported drinking about the same as usual, 25.7% (24.8-26.6%) reported drinking less than usual, and 26.2% (25.4-27.1%) reported drinking more than usual over the past week. Drinking less than usual was independently associated with being younger (OR=0.88 [95% CI=0.83-0.93]), male (OR=0.76 [0.68-0.84]), of an ethnic minority (OR=0.76 [0.61-0.97]), low annual household income (OR=0.74 [0.66-0.83]), having diagnosed or suspected COVID-19 (OR=2.04 [1.72-2.41]), adhering to COVID-19 protective behaviours (OR=1.58 [1.08-2.32]), being significantly stressed about becoming seriously ill from COVID-19 (OR=1.26 [1.08-1.48]) and not being a key worker (OR=0.87 [0.76-0.99]). Drinking more than usual was independently associated with being younger (OR=0.73 [0.69-0.78]), female (OR=1.36 [1.22-1.51]), post-16 qualifications (OR=1.21 [1.04-1.40]), high annual household income (OR=1.43 [1.27-1.61]), being significantly stressed about catching (OR=1.22 [1.03-1.45]) or becoming seriously ill from COVID-19 (OR=1.28 [1.10-1.48]), being significantly stressed about finances (OR=1.43 [1.24-1.66]), and having a diagnosed anxiety disorder (OR=1.24 [1.05-1.46]).

**Conclusions:** In a representative sample of adults in the UK, about half of drinkers reported drinking the same amount of alcohol as usual during the start of the COVID-19 related lockdown, with a quarter drinking more and a quarter drinking less than usual. Drinking more than usual was associated with being younger, female, high socioeconomic position, having an anxiety disorder, and being stressed about finances or COVID-19. These groups may benefit targeted alcohol reduction support if there are further periods of lockdown.

**Registration:** The analysis plan was pre-registered on Open Science Framework (https://osf.io/pnrhq/).

## Introduction

COVID-19 is a respiratory disease caused by the SARS-CoV-2 virus that reached pandemic levels in March 2020 and was declared a Public Health Emergency of International Concern by the World Health Organisation. Reducing alcohol consumption remains a public health priority [1], and public health interventions have been highlighted as one way to “reduce the baseline” demand for acute healthcare services during the pandemic [2]. Alcohol consumption is a leading risk factor for disease burden and the risk of all-cause mortality increases with increasing levels of consumption [3]; 10.8 million adults in England drink at levels that pose some risk to their health [4]. Establishing the factors associated with changes to usual drinking and heaviness of drinking during social distancing and lockdown in the UK is important in order to tailor alcohol interventions to the COVID-19 context and target high-risk groups. This will also be particularly important if there is a second wave of COVID-19 and a return to lockdown.

The UK response to the COVID-19 pandemic required significant disruption to people’s lives. Government advice to practise social distancing came into effect on 16^th^ March and behavioural restrictions (i.e. ‘lockdown’) enforceable by law were introduced on 23^rd^ March. Many businesses, including pubs, bars and restaurants were temporarily closed, and the public were advised to stay at home, avoid unnecessary social contact, and only leave their home for essential journeys and exercise. Early data have indicated that the COVID-19 pandemic has had an impact on people’s alcohol consumption. A large, representative study of adults (n>20,000) in England has shown a substantial increase in the prevalence of high-risk drinking since the lockdown (from 25.1% between April 2019 and February 2020 to 38.3% in April 2020) alongside an increase in attempts to reduce alcohol consumption among high-risk drinkers (from 15.3% to 28.5%) [5]. Other individual-level data suggest that the UK lockdown appeared to have a polarising effect on alcohol consumption [6,7]. A survey of ∼2,000 UK adults commissioned by Alcohol Change UK conducted in early April found that 42% of drinkers had drunk less in the last two weeks compared with 13% who had drunk more [7]. However, decreases in drinking were more likely among lighter drinkers and increases in drinking more likely among heavier drinkers [7]. Generally, data are showing that between a fifth and a third of people in the UK have reported drinking more during lockdown, and that the proportion of people drinking less was often similar to or exceeding the proportion drinking more [6,8].

Collectively, these studies provide evidence that there have been changes in drinking behaviour since lockdown was introduced in the UK. However, there is currently limited evidence on what the risk factors that predict drinking behaviours during the pandemic are: what the socio-demographic risk factors are, but also whether there are any COVID-19 related risk factors, such as being a key worker or diagnosed COVID-19 [8]. It is now critically important to understand who is drinking heavily or drinking more, and why, in order to effectively target support. There are a number of possible reasons people may be drinking more. The lockdown has resulted in people experiencing higher than normal levels of stress relating to social isolation, employment, finances, caring responsibilities, and concerns about catching or becoming ill from COVID-19 [9]. Stress is an important risk factor for the onset and maintenance of alcohol misuse [10,11], though research has also shown it can have a polarising effect on drinking whereby people are more likely to either abstain or drink heavily under stressful conditions, rather than drink lightly or moderately [11]. People who were drinking at harmful levels may develop alcohol dependence triggered by bereavement, job insecurity or troubled relationships during lockdown [12]. People may also drink more heavily to cope with boredom. A recent survey of ∼2,000 adults in the UK found that 35% of furloughed workers were drinking more since lockdown compared with the population average of 24% [13]. It is also interesting to know who and why some people may be drinking less. Reasons people may be drinking less include concerns for health or finances in the context of the pandemic. This is supported by a survey in the UK which found that 89% of people had concerns about their long-term health and 66% reported making changes to protect their long-term health [6]. Previous research has also found that health and financial concerns were the most common reasons for heavy drinkers to consider reducing their drinking [14].

In summary, there is a need for population-based evidence on the factors associated with changes to usual drinking and heaviness of drinking during COVID-19 related social distancing and lockdown in the UK. This study used cross-sectional data from a large well-phenotyped study of adults during the first few weeks of social distancing and lockdown in the UK to answer the following research questions:

1. Among drinkers, are perceived changes to usual drinking over the past week (more versus same as usual, and less versus same as usual) associated with sociodemographic characteristics, diagnosed or suspected COVID-19, adherence to COVID-19 protective behaviours, significant stress about catching or becoming seriously ill from COVID-19, significant stress about finances, significant stress about boredom, recent drop in household income, key worker status, or health conditions?
2. Is heaviness of drinking in the past week associated with sociodemographic characteristics, diagnosed or suspected COVID-19, adherence to COVID-19 protective behaviours, significant stress about catching or becoming seriously ill from COVID-19, significant stress about finances, significant stress about boredom, recent drop in household income, key worker status, or health conditions?

## Methods

### Design and study population

We carried out a cross-sectional analysis of baseline survey data from the UCL COVID-19 Social Study assessing alcohol consumption and measures relating to COVID-19 in the UK for participants recruited between 21^st^ March and 4^th^ April 2020.

The UCL COVID-19 Social Study is a large panel study of the psychological and social experiences of adults (aged 18+) in the UK during the COVID-19 pandemic. The study involves online weekly data collection from participants for the duration of the pandemic in the UK. Data collection began on 21^st^ March 2020 and recruitment is ongoing. In this study, we aggregated the baseline data collected daily for the first two weeks through to 4^th^ April 2020. This study period was chosen as it was at the beginning of social distancing and lockdown in the UK, and therefore we assumed that when participants refer to questions about ‘usual drinking’ that they meant before social distancing measures were introduced. The baseline survey from the study assesses a range of factors including sociodemographics, health, alcohol consumption, confirmed and suspected COVID-19, and behaviours and attitudes relating to COVID-19.

The study contains a well-stratified sample but is not random. Participants were recruited using three primary approaches. First, snowballing was used, including promoting the study through existing networks and mailing lists (including large databases of adults who had previously consented to be involved in health research across the UK), print and digital media coverage, and social media. Second, more targeted recruitment was undertaken focusing on (i) individuals from a low-income background, (ii) individuals with no or few educational qualifications, and (iii) individuals who were unemployed. Third, the study was promoted via partnerships with third sector organisations to vulnerable groups, including adults with pre-existing mental health conditions, older adults, carers, and people experiencing domestic violence or abuse. To account for the non-random nature of the sample, all data are weighted to the proportions of sex, age, ethnicity, education and country of living obtained from the Office for National Statistics [15]. For full details on the recruitment strategies and weighting for the study, visit www.covidsocialstudy.org.

### Measures

#### Changes in drinking over the past week

Changes in drinking over the past week during social distancing and lockdown was measured with the question: “Over the past week have you drunk alcohol more than usual?” with response options: a) less than usual; b) about the same; c) more than usual; and d) I don’t drink alcohol. Only drinkers (i.e. responses a, b or c) were included in the analysis of changes in drinking. For analysis of drinking less or more than usual, those who responded ‘b’ were coded 0 and those who responded ‘a’ or ‘b’, respectively, were coded 1.

#### Heaviness of drinking in the past week

Heaviness of drinking in the past week was assessed using the question: “How many alcoholic drinks have you had in the past week (e.g. how many glasses of wine / pints of beer or cider / shots of spirits)?” with continuous response options from ‘0’ through to ‘21+’.

#### Sociodemographic characteristics

Sociodemographic characteristics included were: age (continuous in years), sex (% female), ethnicity (% white), level of education (% with post-16 qualifications), and annual household income (% >£30,000 [16]). Level of education and income were used as measures of socioeconomic position (SEP). Level of education provides a reliable indication of SEP prior to COVID-19 as it is not affected by recent job loss or furlough, and income gives a strong indication of the economic resources available to the participant and can influence a wide range of circumstances with direct implications for health [18].

#### Diagnosed or suspected COVID-19

##### Participants were asked

“Have you had COVID-19 (coronavirus)?” with response options a) yes diagnosed and recovered, b) yes diagnosed and still ill, c) not formally diagnosed but suspected, d) no. Diagnosed or suspected COVID-19 was coded 1 for those who responded ‘a’, ‘b’ or ‘c’ and 0 for those who responded ‘d’.

#### Adherence to COVID-19 protective behaviours

##### Adherence to COVID-19 protective behaviours was assessed with the question

“Are you following the recommendation from authorities to prevent spread of COVID-19?” with responses on a scale from 1 (not at all) to 7 (very much so). Responses of 5 and above were coded 1 (indicating adherence) and responses of 4 and below were coded 0.

#### Stress about COVID-19, finances or boredom

##### Stress about COVID-19, finances or boredom was assessed with the question

“Have any of these things been causing you significant stress? (e.g. they have been constantly on your mind or have been keeping you awake at night)”. Response options included i) catching COVID-19, ii) becoming seriously ill from COVID-19, iii) finances, and iv) boredom. For each of these four response options, a variable was created where those who reported stress about the relevant outcome were coded 1, else they were coded 0.

#### Recent drop in household income

Recent drop in household income was assessed with the question “Have you experienced any of the following in the past week?”. Those who reported ‘lost your job/been unable to do paid work’, ‘your spouse/partner lost their job or was unable to do paid work’ or ‘major cut in household income (e.g. due to you or your partner being furloughed/put on leave/not receiving sufficient work)’ were coded 1 for recent drop in household income and those who reported none of these were coded 0.

#### Key worker status

Key worker status may be associated with increased exposure to COVID-19. Participants were asked: “Are you currently fulfilling any of the government’s identified ‘key worker’ roles?” Participants were coded 1 if they selected any of the ‘key worker’ roles (e.g. health care work, teacher, food chain worker) and those who responded ‘none of these’ were coded 0.

#### Health conditions

The presence of health conditions was assessed with the question: “Do you have any of the following medical conditions?” Those who selected ‘high blood pressure’, ‘diabetes’, ‘heart disease’, ‘lung disease (e.g. asthma or COPD)’, or ‘cancer’ were coded 1 and those who selected none of these were coded 0. The presence of anxiety disorders was assessed with the same question, with those who selected ‘clinically-diagnosed anxiety’ coded 1 and those who did not select this response coded 0.

#### Survey date

Survey date was coded 0 for those surveys completed on 21^st^, 22^nd^ or 23^rd^ March (i.e. prior to lockdown commencing), and subsequent dates were coded 1, 2, 3,…, n (i.e. reflecting the progression of time from the start of lockdown).

Full details of all of the measures is available in Supplementary File 1.

### Analyses

The protocol and analysis plan was pre-registered on Open Science Framework (https://osf.io/pnrhq/). Analyses were conducted using R studio on complete cases. To account for the non-random nature of the sample, all data were weighted to the proportions of sex, age, ethnicity, education, and country of living obtained from the Office for National Statistics [15]. Unweighted analyses are presented in supplementary material.

The proportion (and 95% confidence interval [CI]) of those who reported drinking more than usual, about the same as usual, and less than usual over the past week was calculated. The mean (and standard deviation [SD]) of heaviness of drinking in the past week was calculated.

Generalized linear modeling (logistic or linear, as appropriate) was used to examine unadjusted and adjusted associations of associations between i) drinking less than usual (versus about the same as usual), ii) drinking more than usual (versus about the same as usual), and iii) heaviness of drinking with the following: age, sex, ethnicity, education level, income, diagnosed or suspected COVID-19, adherence to COVID-19 protective behaviours, significant stress about catching COVID-19, significant stress about becoming seriously ill from COVID-19, significant stress about finances, significant stress about boredom, recent drop in household income, key worker status, presence of health conditions and anxiety disorder, and survey date. The continuous variable age was transformed by dividing the original variable by the SD to express the variable as a proportion of the SD for ease of interpretation of the odds ratio. Sensitivity analyses were conducted for the associations with heaviness of drinking which repeated the unadjusted and adjusted linear regression models excluding the response option ‘21+’ as this is a non-linear response option.

## Results

### Participant characteristics

A total of 33,644 participants responded to the survey between 21^st^ March and 4^th^ April 2020, of whom 30,375 (90.3%; weighted n=30,516) provided complete data on the variables included in the present analyses. Of the analysed sample, 22,113 were drinkers (65.7%, weighted n=21,212). Participant characteristics are shown in Table 1 (unweighted data are shown in Supplementary Table 1). The mean age was 47.8 years; around half were female (49.6%) and had an annual household income over £30,000 (52.9%); and the majority were white (88.0%) and had post-16 education qualifications (69.3%).

**Table 1:**
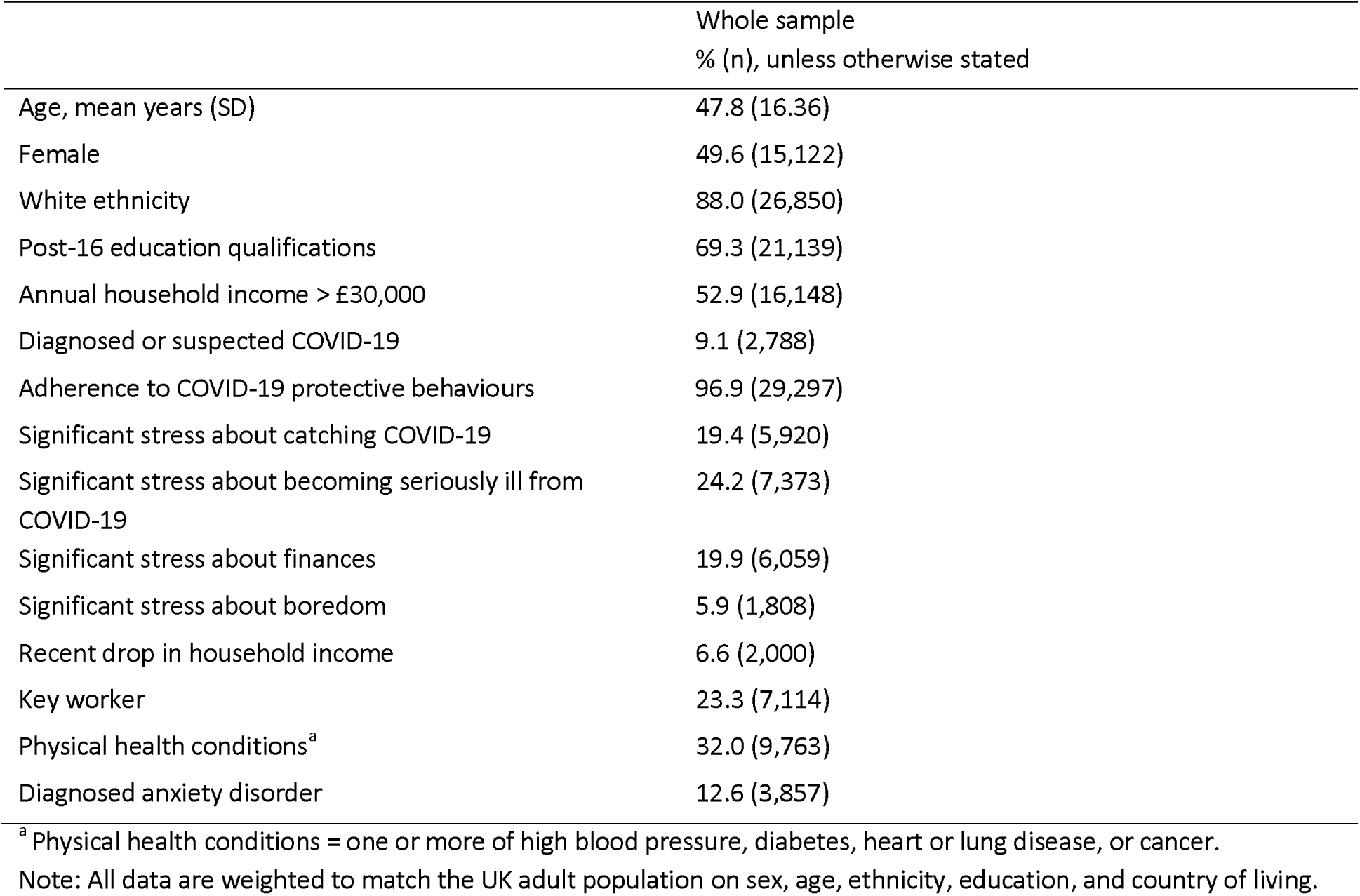
Participant characteristics

### Associations with changes in drinking over the past week

Of the 22,113 drinkers (weighted n=21,212), 48.1% (95% CI=47.0-49.1%; weighted n =10,196) reported drinking about the same as usual, 25.7% (95% CI=24.8-26.6%; weighted n =5,451) reported drinking less than usual, and 26.2% (95% CI=25.4-27.1%; weighted n =5,566) reported drinking more than usual over the past week.

Drinking less than usual, compared with the same as usual, over the past week was independently associated with being younger, male, an ethnic minority, having an annual household income of less than £30,000, having diagnosed or suspected COVID-19, adhering to COVID-19 protective behaviours, being significantly stressed about becoming seriously ill from COVID-19, and not being a key worker (Table 2).

**Table 2:** Factors associated with drinking less than usual, compared with same as usual, over the past week

|  | Proportion drinking<br>less than usual, % (n) | Unadjusted<br>OR (95% CI) | p | Adjusted<sup>b</sup><br>OR<sub>adj</sub> (95% CI) | p |
| --- | --- | --- | --- | --- | --- |
| Age, in years | - | 0.90 (0.86, 0.95) | <.001 | 0.88 (0.83, 0.93) | <.001 |
| Sex: Male | 36.5 (3,249) |  |  |  |  |
| Female | 32.7 (2,201) | 0.85 (0.77, 0.93) | <.001 | 0.76 (0.68, 0.84) | <.001 |
| Ethnicity: Ethnic minority | 41.7 (680) |  |  |  |  |
| White | 34.0 (4,771) | 0.72 (0.57, 0.91) | .005 | 0.76 (0.61, 0.97) | .028 |
| Post-16 education<br>qualifications: No | 36.5 (1,718) |  |  |  |  |
| Yes | 34.1 (3,733) | 0.90 (0.79, 1.02) | .101 | 0.90 (0.79, 1.03) | .135 |
| Annual household income: <<br>£30,000 | 38.0 (2,655) |  |  |  |  |
| > £30,000 | 32.2 (2,796) | 0.78 (0.70, 0.86) | <.001 | 0.74 (0.66, 0.83) | <.001 |
| Diagnosed or suspected<br>COVID-19: No | 33.1 (4,686) |  |  |  |  |
| Yes | 50.9 (765) | 2.09 (1.76, 2.48) | <.001 | 2.04 (1.72, 2.41) | <.001 |
| Adherence to COVID-19 protective behaviours: No | 28.6 (172) |  |  |  |  |
| Yes | 35.1 (5,279) | 1.35 (0.94, 1.94) | .103 | 1.58 (1.08, 2.32) | .020 |
| Significant stress about catching COVID-19: No | 34.1 (4,490) |  |  |  |  |
| Yes | 39.1 (961) | 1.24 (1.07, 1.44) | .004 | 1.01 (0.84, 1.21) | .941 |
| Significant stress about becoming seriously ill from COVID-19: No | 33.4 (4,175) |  |  |  |  |
| Yes | 40.6 (1,276) | 1.36 (1.20, 1.55) | <.001 | 1.26 (1.08, 1.48) | .004 |
| Significant stress about finances: No | 34.0 (4,468) |  |  |  |  |
| Yes | 39.5 (984) | 1.27 (1.09, 1.47) | .002 | 1.02 (0.87, 1.19) | .829 |
| Significant stress about boredom: No | 34.4 (5,109) |  |  |  |  |
| Yes | 43.5 (342) | 1.47 (1.12, 1.92) | .005 | 1.19 (0.91, 1.57) | .206 |
| Recent drop in household income: No | 34.4 (5,076) |  |  |  |  |
| Yes | 42.0 (375) | 1.38 (1.10, 1.72) | .005 | 1.17 (0.93, 1.48) | .184 |
| Key worker: No | 35.3 (4,255) |  |  |  |  |
| Yes | 33.2 (1,196) | 0.91 (0.80, 1.03) | .144 | 0.87 (0.76, 0.99) | .030 |
| Physical health conditions<sup>a</sup>: No | 34.5 (3,669) |  |  |  |  |
| Yes | 35.5 (1,782) | 1.04 (0.93, 1.17) | .464 | 1.07 (0.94, 1.21) | .312 |
| Diagnosed anxiety disorder: No | 34.2 (4,864) |  |  |  |  |
| Yes | 40.7 (587) | 1.32 (1.10, 1.57) | .002 | 1.10 (0.92, 1.32) | .316 |
| Survey date | - | 1.02 (1.00, 1.04) | .051 | 1.01 (0.99, 1.03) | .204 |

<sup>a</sup> Physical health conditions = high blood pressure, diabetes, heart or lung disease, or cancer.
<sup>b</sup> Multivariable model fully adjusted for all variables in the table.
Note: All data are weighted to match the UK adult population on sex, age, ethnicity, education, and country of living.

Drinking more than usual, compared with the same as usual, over the past week was independently associated with being younger, female, having post-16 education qualifications, having an annual household income over £30,000, being significantly stressed about catching or becoming seriously ill from COVID-19, being significantly stressed about finances, having a diagnosed anxiety disorder, and with survey date (Table 3).

**Table 3:** Factors associated with drinking more than usual, compared with same as usual, over the past week

|  | Proportion drinking more than usual, % (n) | Unadjusted OR (95% CI) | p | Adjusted<sup>b</sup> OR<sub>adj</sub> (95% CI) | p |
| --- | --- | --- | --- | --- | --- |
| Age, in years | - | 0.66 (0.63, 0.69) | <.001 | 0.73 (0.69, 0.78) | <.001 |
| Sex: Male | 30.1 (2,443) |  |  |  |  |
| Female | 40.8 (3,123) | 1.60 (1.44, 1.76) | <.001 | 1.36 (1.22, 1.51) | <.001 |
| Ethnicity: Ethnic minority | 37.1 (561) |  |  |  |  |
| White | 35.1 (5,004) | 0.92 (0.73, 1.15) | .450 | 1.08 (0.84, 1.39) | .555 |
| Post-16 education qualifications: No | 28.6 (1,194) |  |  |  |  |
| Yes | 37.7 (4,372) | 1.51 (1.32, 1.73) | <.001 | 1.21 (1.04, 1.40) | .012 |
| Annual household income: < £30,000 | 29.2 (1,788) |  |  |  |  |
| > £30,000 | 39.2 (3,778) | 1.56 (1.40, 1.74) | <.001 | 1.43 (1.27, 1.61) | <.001 |
| Diagnosed or suspected COVID-19: No | 34.8 (5,053) |  |  |  |  |
| Yes | 40.9 (512) | 1.30 (1.10, 1.54) | .003 | 1.06 (0.89, 1.26) | .517 |
| Adherence to COVID-19 protective behaviours: No | 34.2 (223) |  |  |  |  |
| Yes | 35.4 (5,342) | 1.05 (0.75, 1.48) | .777 | 0.89 (0.61, 1.30) | .553 |
| Significant stress about catching COVID-19: No | 33.6 (4,405) |  |  |  |  |
| Yes | 43.6 (1,161) | 1.53 (1.34, 1.74) | <.001 | 1.22 (1.03, 1.45) | .020 |
| Significant stress about becoming seriously ill from COVID-19: No | 33.1 (4,120) |  |  |  |  |
| Yes | 43.6 (1,446) | 1.56 (1.40, 1.75) | <.001 | 1.28 (1.10, 1.48) | <.001 |
| Significant stress about finances: No | 33.2 (4,312) |  |  |  |  |
| Yes | 45.4 (1,254) | 1.67 (1.47, 1.90) | <.001 | 1.43 (1.24, 1.66) | <.001 |
| Significant stress about boredom: No | 34.8 (5,215) |  |  |  |  |
| Yes | 44.1 (351) | 1.47 (1.15, 1.89) | .002 | 1.10 (0.84, 1.44) | .489 |
| Recent drop in household income: No | 34.3 (5,060) |  |  |  |  |
| Yes | 49.4 (506) | 1.87 (1.54, 2.26) | <.001 | 1.23 (1.00, 1.51) | .055 |
| Key worker: No | 34.6 (4,128) |  |  |  |  |
| Yes | 37.4 (1,437) | 1.13 (1.01, 1.26) | .032 | 0.92 (0.82, 1.03) | .191 |
| Physical health conditions<sup>a</sup>: No | 37.8 (4,237) |  |  |  |  |
| Yes | 29.1 (1,328) | 0.67 (0.60, 0.75) | <.001 | 0.91 (0.80, 1.03) | .128 |
| Diagnosed anxiety disorder: No | 34.0 (4,819) |  |  |  |  |
| Yes | 46.6 (747) | 1.69 (1.45, 1.96) | <.001 | 1.24 (1.05, 1.46) | .011 |
| Survey date | - | 1.07 (1.06, 1.09) | <.001 | 1.06 (1.04, 1.08) | <.001 |

<sup>a</sup> Physical health conditions = high blood pressure, diabetes, heart or lung disease, or cancer.
<sup>b</sup> Multivariable model fully adjusted for all variables in the table.
Note: All data are weighted to match the UK adult population on sex, age, ethnicity, education, and country of living.

Unweighted analyses are reported in Supplementary Table 2 and Supplementary Table 3 and the pattern of results is broadly similar.

### Associations with heaviness of drinking in the past week

The mean heaviness of drinking in the past week was 4.0 alcoholic drinks (SD=5.34). Heaviness of drinking in the past week was independently positively associated with being older, male, white ethnicity, having post-16 education qualifications, having an annual household income over £30,000, not adhering to COVID-19 protective behaviours, not being significantly stressed about catching COVID-19, a recent drop in household income, not being a key worker, not having physical health conditions or a diagnosed anxiety disorder, and with survey date (Table 4).

**Table 4:** Factors associated with heaviness of drinking in the past week

|  | Mean number of drinks in past week (SD) | Unadjusted B (95% CI) | p | Adjusted<sup>b</sup> B<sub>adj</sub> (95% CI) | p |
| --- | --- | --- | --- | --- | --- |
| Age, in years | - | 0.59 (0.50, 0.68) | <.001 | 0.64 (0.54, 0.75) | <.001 |
| Sex: Male | 4.8 (5.92) |  |  |  |  |
| Female | 3.2 (4.54) | -1.61 (-1.80, -1.42) | <.001 | -1.29 (-1.47, -1.10) | <.001 |
| Ethnicity: Ethnic minority | 2.3 (3.97) |  |  |  |  |
| White | 4.2 (5.46) | 1.85 (1.54, 2.15) | <.001 | 1.67 (1.36, 1.98) | <.001 |
| Post-16 education qualifications: No | 3.6 (5.30) |  |  |  |  |
| Yes | 4.2 (5.35) | 0.59 (0.36, 0.82) | <.001 | 0.61 (0.38, 0.85) | <.001 |
| Annual household income: < £30,000 | 3.2 (5.04) |  |  |  |  |
| > £30,000 | 4.7 (5.50) | 1.46 (1.27, 1.65) | <.001 | 1.53 (1.32, 1.73) | <.001 |
| Diagnosed or suspected COVID-19: No | 4.0 (5.37) |  |  |  |  |
| Yes | 3.6 (5.03) | -0.36 (-0.66, -0.05) | .023 | -0.10 (-0.40, 0.20) | .516 |
| Adherence to COVID-19 protective behaviours: No | 4.6 (6.50) |  |  |  |  |
| Yes | 3.9 (5.28) | -0.64 (-1.36, 0.08) | .083 | -0.96 (-1.65, -0.27) | .006 |
| Significant stress about catching COVID-19: No | 4.2 (5.40) |  |  |  |  |
| Yes | 3.2 (5.00) | -0.97 (-1.20, -0.74) | <.001 | -0.31 (-0.58, -0.03) | .029 |
| Significant stress about becoming seriously ill from COVID-19: No | 4.2 (5.42) |  |  |  |  |
| Yes | 3.3 (5.01) | -0.91 (-1.12, -0.71) | <.001 | -0.21 (-0.45, 0.03) | .084 |
| Significant stress about finances: No | 4.1 (5.37) |  |  |  |  |
| Yes | 3.3 (5.18) | -0.79 (-1.03, -0.55) | <.001 | -0.01 (-0.26, 0.23) | .916 |
| Significant stress about boredom: No | 4.0 (5.34) |  |  |  |  |
| Yes | 3.6 (5.37) | -0.43 (-0.87, 0.02) | .060 | 0.23 (-0.20, 0.66) | .286 |
| Recent drop in household income: No | 4.0 (5.34) |  |  |  |  |
| Yes | 4.0 (5.40) | 0.01 (-0.38, 0.40) | .967 | 0.44 (0.03, 0.84) | .033 |
| Key worker: No | 4.1 (5.48) |  |  |  |  |
| Yes | 3.6 (4.85) | -0.47 (-0.68, -0.27) | <.001 | -0.32 (-0.52, -0.12) | .002 |
| Physical health conditions<sup>a</sup>: No | 4.0 (5.19) |  |  |  |  |
| Yes | 4.0 (5.65) | -0.003 (-0.22, 0.21) | .981 | -0.33 (-0.56, -0.10) | .0049 |
| Diagnosed anxiety disorder: No | 4.2 (5.39) |  |  |  |  |
| Yes | 2.7 (4.78) | -1.44 (-1.68, -1.20) | <.001 | -0.52 (-0.76, -0.28) | <.001 |
| Survey date | - | 0.01 (-0.02, 0.04) | .572 | 0.04 (0.0003, 0.07) | .048 |

<sup>a</sup> Physical health conditions = high blood pressure, diabetes, heart or lung disease, or cancer.
<sup>b</sup> Multivariable model fully adjusted for all variables in the table.
Note: All data are weighted to match the UK adult population on sex, age, ethnicity, education, and country of living.

Sensitivity analyses were conducted in which the non-linear response option ‘21+’ was excluded (reported in Supplementary Table 4) and the pattern of results remained similar. Unweighted analyses are reported in Supplementary Table 5 and the pattern of results is broadly similar.

## Discussion

Among a nationally representative sample of 33,644 adults in the UK surveyed in the first two weeks of national COVID-19 related social distancing and lockdown restrictions, adults drank a mean of 4 alcoholic drinks in the past week with almost a third not drinking (34.3%). Among people who drank, 48% reported drinking about the same as usual, 26% reported drinking more and 26% reported drinking less than usual over the past week. These findings are in line with those from other recent surveys where between a fifth and a third reported drinking more during lockdown, and that the proportion of people drinking less was often similar to or exceeding the proportion drinking more [8].

Younger drinkers were more likely than older drinkers to report changes in their drinking behaviour during lockdown (both drinking less and drinking more over the past week), which is consistent with previous findings [19,20]. Younger people in the UK may be drinking more because they have been most affected by lockdown with large declines in mental well-being being seen [21]. Younger people have also reported worries about the impact of lockdown on university courses, work and relationships [22]; and during lockdown, loneliness levels have been the highest and life satisfaction levels the lowest among younger adults [23]. Changes in the opposite direction (i.e. some younger people drinking less than usual) may be attributable to the closure of on-trade drinking locations (e.g. bars, pubs, restaurants, hotels) during the beginning of lockdown as on-trade drinking is more common among younger drinkers [24]. This would fit with the finding from the Global Drugs Survey (where young people are over-represented) that less exposure to settings and people associated with drinking was a key reason for reductions in drinking [25]. Closures of on-trade drinking locations may have affected older people less (as they tend towards off-trade drinking [24]) and this could explain why their drinking remains relatively unaffected. However, despite older people being less likely to have changed their drinking, their heaviness of drinking was greater than younger people at the start of lockdown, which may be because ‘drinking at home alone’ is more common among older people [24]. Therefore, it is important that older drinkers are not overlooked in terms of alcohol reduction support during lockdown.

Women were more likely to report drinking more than usual and men were more likely to report drinking less than usual during lockdown. The latter finding is consistent with two previous surveys [19,20]. Women may be drinking more than men during lockdown because they have been more negatively affected by lockdown. Lockdown appears to have increased gender inequalities in both paid and unpaid work, with women more likely than men to lose their jobs and carry the burdens of increased childcare and housework [26,27]. There has also been a substantial negative effect of lockdown on mental well-being, which can lead to increased drinking. This negative effect on mental well-being is largely driven by women [21,28] due to family and caring responsibilities, and social factors such as increased loneliness [21]. Despite women being more likely to report drinking more than usual, heaviness of drinking in the past week was positively associated with being male. This may be because ‘drinking at home alone’ is more common among men [24]. However, it is important to note that this was also the case before lockdown began and that despite a greater increase in women than men drinking at high risk in England during lockdown, the overall number is still greater for men [29].

Drinkers of a low SEP (indicated by income) were more likely to be drinking less than usual. Those of high SEP (indicated by income and education qualifications) were more likely to be drinking more than usual over the past week, and more heavily. Previous surveys have also found that a greater proportion of those of high SEP reported drinking more during lockdown than those of low SEP [19], and that the proportion drinking on five or more days a week after lockdown increased most among those of high SEP [30]. All of these findings taken together suggest that there is a polarisation based on SEP in terms of how drinking behaviour has changed during lockdown. It is reassuring that these findings suggest that changes in drinking behaviour during lockdown have not widened existing health inequalities, as those of low SEP are more at risk of alcohol-related harm for the same level of consumption compared with those of high SEP [31].

Those of an ethnic minority were more likely than those of white ethnicity to be drinking less than usual and being of white ethnicity was positively associated with heaviness of drinking. There is clear evidence that COVID-19 does not affect all groups equally and minority ethnic groups have a greater risk of infection, experience more severe symptoms and higher rates of death [32,33]. Therefore, minority ethnic groups being more likely to drink less than usual may be a response to more general health concerns in the context of the pandemic and protecting their long-term health by reducing their alcohol consumption [6,14]. Similarly, those who had had diagnosed or suspected COVID-19 or were adhering to COVID-19 protective behaviours were more likely to report drinking less than usual over the past week, and adhering to protective behaviours was also negatively associated with heaviness of drinking. These groups are more likely to have COVID-19-related health concerns and therefore may be taking steps to protect their health by drinking less. Those who were not a key worker were more likely to be drinking less than usual, though not being a key worker was also positively associated with the heaviness of drinking.

We also found that psychological factors predicted changes in drinking behaviours. Stress about catching COVID-19, becoming seriously ill from COVID-19 and finances were all associated with drinking more than usual over the past week, and stress about catching COVID-19 and experiencing a recent drop in household income were also positively associated with heaviness of drinking. This is in line with previous research showing that stress is a risk factor for the onset of alcohol misuse [10,11]. However, stress about becoming seriously ill from COVID-19 was also associated with drinking less than usual over the past week. This may be due to health and economic stress in the context of the pandemic [6,14]; stress likely has a polarising effect whereby it motivates some people to cut down to improve health and others to drink heavily as a maladaptive coping strategy [11].

Exploring mental health more broadly, having a diagnosed anxiety disorder was associated with drinking more than usual though was negatively associated with heaviness of drinking, along with having a physical health condition. It is possible that people with anxiety disorders are changing their drinking behaviour to self-medicate [34] or as a coping mechanism [35] during a period of increased anxiety because of the pandemic. This finding fits with that of the Global Drugs Survey where reasons for increased drinking during lockdown were commonly reported as being due to worry and depression [25]. This may indicate that people are drinking as a maladaptive coping mechanism and future research should investigate the motives behind changes in drinking behaviour among people with mental health conditions.

### Implications

This study has important implications in terms of groups that may need targeted support for alcohol reduction to counteract an increase in drinking during the start of the COVID-19 related lockdown. With one in four drinkers reporting an increase in consumption since lockdown began, these findings emphasise the need for alcohol reduction support with a focus on those groups who are drinking more than usual and drinking heavily. This may be particularly important if a second wave of COVID-19 results in another lockdown. However, more research is needed to understand whether these changes have been sustained or whether people’s drinking have returned (or will return) to usual levels as they gradually resume their normal lives. Heaviness of drinking was positively associated with survey date indicating that heaviness of drinking increased with the time spent in lockdown, therefore it’s important to establish whether any initial changes during the start of lockdown continued, or possibly were even exacerbated.

### Strengths and limitations

A key strength of this study was the large sample size and broad range of measures collected, permitting this detailed analysis of potential factors predicting drinking behaviour during the start of social distancing and lockdown in the UK. The collection of data in real time while the pandemic was at its peak was also a strength, minimising potential recall bias that is likely to be present in any future studies that collect data retrospectively. Another major strength of this study was being able to assess how sociodemographic characteristics and COVID-19-related impacts, such as key worker status, were associated with changes in alcohol consumption – an important area of research highlighted in a recent report by the Institute of Alcohol Studies [8].

A limitation of this study relates to the alcohol consumption measures used. The measure of changes in drinking over the past week did not distinguish between changes in frequency of consumption, quantity per occasion or heavy episodic drinking, which is an important topic for future research. Furthermore, the changes in drinking measure used the reference ‘usual drinking’ rather than specifying before lockdown. Though the study period was chosen as the start of social distancing and lockdown in the UK and therefore we have assumed that participants interpreted ‘usual’ as meaning before lockdown restrictions were introduced in the UK. The measure of heaviness of drinking asks about the number of alcoholic drinks consumed and therefore the overall index does not directly correspond with standardised units. As the survey was conducted at the start of lockdown and the heaviness of drinking measure refers to the past week, the associations detected may reflect the characteristics of people who were drinking alcohol more heavily prior to the COVID-19 pandemic rather than as a response to the lockdown in the UK. However, it was still important to consider the heaviness of drinking as this gives an important indication for the public health implications of respondents’ alcohol consumption.

The study did not include a measure of baseline alcohol consumption, although it did ask about consumption in the previous week, which occurred before lockdown came in. Future studies with prior data may be able to ascertain how long-term trajectories of alcohol consumption were affected by the pandemic. Whilst we do not know the prevalence of high-risk drinking in this sample, our finding that 26% of adults in the UK reported drinking more could be reconciled with data on the change in high-risk drinking prevalence in England [5], particularly if those who reported drinking more were borderline high-risk drinkers before lockdown.

## Conclusions

About half of adult drinkers in the UK reported drinking about the same as usual, a quarter reported drinking more than usual, and a quarter reported drinking less than usual over the past week during the start of the COVID-19 related lockdown. Drinking more than usual during the start of lockdown was associated with being younger, female, of high SEP, having an anxiety disorder, being stressed about finances, or being stressed about catching or becoming seriously ill from COVID-19. This highlights that there are certain groups who need alcohol reduction support during lockdown and may be particularly important to target support for if there is a second wave of COVID-19.

## Data Availability

Anonymous data will be made publicly available after the end of the pandemic.

## Governance and ethics

Ethical approval for the COVID-19 Social Study was granted by the UCL Ethics Committee [12467/005]. All participants provided fully informed consent. The study is GDPR compliant.

## Acknowledgements

The Covid-19 Social Study was funded by the Nuffield Foundation [WEL/FR-000022583], but the views expressed are those of the authors and not necessarily the Foundation. The study was also supported by the MARCH Mental Health Network funded by the Cross-Disciplinary Mental Health Network Plus initiative supported by UK Research and Innovation [ES/S002588/1], and by the Wellcome Trust [221400/Z/20/Z]. DF was funded by the Wellcome Trust [205407/Z/16/Z]. CG is funded by CRUK [C1417/A22962] and NIHR. SJ and JB are funded by CRUK [C1417/A22962]. MO is funded by NIHR.

The researchers are grateful for the support of a number of organisations with their recruitment efforts including: the UKRI Mental Health Networks, Find Out Now, UCL BioResource, SEO Works, FieldworkHub, and Optimal Workshop. The study was also supported by HealthWise Wales, the Health and Car Research Wales initiative, which is led by Cardiff University in collaboration with SAIL, Swansea University. The funders had no final role in the study design; in the collection, analysis and interpretation of data; in the writing of the report; or in the decision to submit the paper for publication. All researchers listed as authors are independent from the funders and all final decisions about the research were taken by the investigators and were unrestricted.

## Notes

### Competing Interest Statement

CG, SJ, MO, AS and DF declare no conflicts of interest. JB has received unrestricted research grants from Pfizer related to smoking cessation.

### Clinical Protocols

https://osf.io/pnrhq/

